# Epigenetics for Public Consumption: Evaluating Science Communication Strategies and Practices on YouTube

**DOI:** 10.64898/2026.08.14.26360379

**Authors:** Aantaki Raisa, Irania Santaliz Moreno, Amy Ayala, Jada G. Hamilton, Amy McQueen, George P. Souroullas, Julia Maki, Erika A. Waters

**Affiliations:** Division of Public Health Sciences, Department of Surgery, Washington University in St. Louis, St. Louis, Missouri, USA; Division of Biology and Biomedical Sciences Washington University in St. Louis, St. Louis, Missouri, USA; Institute for Community Health Innovation, University of Arkansas for Medical Sciences, Northwest, Springdale, AR; Department of Psychiatry & Behavioral Sciences, Memorial Sloan Kettering Cancer Center, New York, New York, USA; School of Public Health, Washington University in St. Louis, St. Louis, Missouri, USA; Division of Oncology, Department of Medicine, Washington University School of Medicine, St. Louis, USA

**Author notes:** **Corresponding author**: Erika A. Waters. Aantaki Raisa, PhD, MA, CRediT: Conceptualization, formal analysis, investigation, methodology, data curation, visualization, writing-original draft, writing-review & editing. Irania Santaliz Moreno, MS, CRediT: formal analysis, investigation, data curation, review and editing. Amy Ayala, CRediT: investigation, review and editing. Jada G. Hamilton, PhD, MPH, CRediT: Conceptualization, methodology, Manuscript review and editing. Amy McQueen, CRediT: Methodology, review and editing. George P. Souroullas, PhD, CRediT: Conceptualization, Manuscript review and editing. Julia Maki, CRediT: Review and editing. Erika A. Waters, PhD, MPH, <u>CRediT</u>: Conceptualization, methodology, funding acquisition, supervision, writing— original draft, writing—review & editing.

**Keywords:** public engagement with science, epigenomics, content analysis, science education, social media

## Abstract

**Background:** Epigenetics—the study of reversible changes in gene expression without altering the underlying DNA sequence—is increasingly applied in medical, commercial, and policy contexts. Yet, little is known about how this emerging science is communicated to the public. The purpose of this study was to examine communication strategies, sources, and modalities in epigenetic-related videos on YouTube- the most accessed platform for informal science education.

**Methods:** We conducted a mixed-methods content analysis of 294 YouTube videos on epigenetics by conducting a keyword-based search on October 17, 2023. Video transcripts and meta-data were coded using a codebook developed both deductively and inductively. Qualitative analysis examined how communication strategies were used within videos and identified emergent themes (RQ1). Quantitative analyses examined the frequency of video and channel characteristics (RQ2), and presentation modalities (RQ3).

**Results:** Findings reveal poor alignment with science communication best practices (RQ1): over 92% of videos failed to acknowledge scientific uncertainty, the comprehensibility level exceeded the recommended 8th-grade level (e.g., average readability grade 10.7), and professional research organizations were notably absent. Narrators were mostly male (56.7%) and white-presenting (73.7%) (RQ2). The majority of the videos used multi-modal strategies (e.g., visual texts mixed with animation and voice-over narration) to communicate epigenetic information (RQ3).

**Conclusion:** Findings highlight the need for professional research organizations to be more proactive in public epigenetic communication efforts. Increasing narrator demographic diversity could broaden audience reach. Evidence-based communication tools are needed for health or science communicators discussing epigenetics on social media.

## Introduction

The field of epigenetics studies stable yet reversible changes in gene expression that occur without altering the underlying DNA sequence ^1^. Advances in epigenetics have significantly influenced biomedical research and practice in recent years ^2^. Discoveries in epigenetics have enhanced our understanding of the mechanisms of cancer, diabetes, and obesity ^3–5^, leading to the development of personalized drugs and therapeutics ^6^.

Moreover, epigenetic findings are also being used to inform health and public policies ^7^, and to sell direct-to-consumer products, such as epigenetics-based skincare products ^8^, epigenetic aging tests ^9^, and disease diagnostics ^10^. These discoveries and applications have increased media coverage of epigenetics ^11^ and, consequently, have generated greater public interest ^12^.

Epigenetics helps explain how our environment ^13^, diet ^14^, stress ^15^, and trauma ^16^ interact with gene expression and disease risk. Consequently, it is increasingly mentioned in public discussions to explain the health effects of lived experiences, such as environmental exposures, lifestyle behaviors, and generational trauma ^17^. Information about epigenetics is now being disseminated by sources beyond scientists and science journalists, including health influencers on social media ^12,18,19^.

Communication about emerging and quickly-evolving scientific subjects like epigenetics must be carefully evaluated for clarity and accuracy ^11,20^. Otherwise, the public may develop an inaccurate understanding of epigenetics, which could lead to maladaptive health beliefs and behaviors or stigma for health conditions ^21^. While numerous communication strategies and recommendations exist for presenting health-related information to the public, it remains unexplored whether these strategies are applied to content that communicates epigenetic information on social media (where much modern public-facing science communication predominantly occurs) ^22^. We investigate this issue by first identifying how YouTube videos about epigenetics align with the recommended best practices in science communication. Then we investigate *who* communicates about epigenetics (its source) and *how* epigenetics is communicated (its modality) in these videos. We focus on YouTube because it was the most popular platform for informal science education at the time this study was conducted ^23,24^.

### Best practices in science communication

Science communication is the exchange of information and viewpoints to foster a greater understanding of a scientific topic ^25^. Best practice recommendations for science communication are common and widely available. As one example, the American Society for Cell Biology suggests: 1) know your audience, 2) avoid jargon, and 3) use metaphors ^26^. The National Academy of Medicine underscores the importance of the communicator— the source of the message — in establishing trust and credibility in scientific communication ^27^. Similarly, the National Science Teaching Association’s “ABCs of science communication” suggests effective modalities (e.g., audiovisuals alongside textual communication) ^28^. While science communication toolkits and general recommendations are intended for general audiences, we are unaware of studies that investigate whether these strategies are applied in YouTube videos about a topic as complex as epigenetics. Therefore, our first research question (RQ) asks: *What communication strategies are used in YouTube videos about epigenetics?* (RQ1)

To identify *who* is communicating epigenetic science on YouTube and *how*, we focus on the following components of communication (as described by Berlo’s Transmission Model of Communication ^29^): 1) channel, 2) receiver, 3) source, and 4) message. In this study, the channel (YouTube) and receiver (the public) remain constant. The source of communication refers to the account that uploaded the video and the video’s narrator. To determine *who* shares epigenetic information, we ask: *What are the sources disseminating epigenetic information on YouTube?* (RQ2)

We further divided the message component into two subcategories: 1) content and 2) modality ^29^. Message content refers to the information the communicator intends their audience to receive ^30^. Prior research has examined the content of epigenetic information communicated in YouTube videos, identifying that while the basic molecular mechanisms are generally portrayed accurately, these videos frequently contain exaggerated, unsubstantiated health claims and overemphasize personal control and omit critical scientific nuances ^19^. Message modality refers to the mode(s) of communication in the videos, such as text or multimodal formats. To identify *how* epigenetic information is communicated, we ask: *What modalities are used in YouTube videos to communicate epigenetic information?* (RQ3).

Together with our prior findings on message content including exaggerated health claims on YouTube about epigenetics, answering these research questions will provide a comprehensive picture of the current state of epigenetic communication on this platform and identify specific areas for improvement in science communication practices.

## Method

We conducted a content analysis of YouTube videos about epigenetics, following the Standards for Reporting Qualitative Research (SRQR) detailed in Appendix 1.

### Preliminary research and initial codebook development

In May 2023, a preliminary search of YouTube videos about epigenetics and existing literature informed the development of a draft codebook for identifying science communication strategies. Initial codes included: *use of metaphors, analogies, or similes*; *acknowledgment of scientific uncertainty; scientific examples;* and *use of undefined jargon*.

### Data collection

On October 17, 2023, we used the web data extraction platform Apify ^31^ to search YouTube with terms: epigenetic, epigenetics, epigenome, epigenomic, and epigenomics. We set limits to collect up to 500 regular YouTube videos and 100 YouTube Shorts^1^ (with a total return of 472 regular videos and 100 Shorts). Data collected for each video included URL, duration, publication date, uploader account name, and video type (regular vs. short). All videos were transcribed using a transcription website ^32^ and reviewed for accuracy, and the transcripts were imported into NVivo Version 12 for coding. Shorts containing only on-screen text (no audio) were manually transcribed.

### Inclusion criteria

Videos were included if they: 1) were ≤10 minutes long (shorter videos better retain attention in informal science education) ^33,34^; 2) were in English; 3) defined/described epigenetics or linked it to health outcomes; 4) contained at least one complete statement about epigenetics; and 5) had audible speech (applied only to regular videos, as speechless Shorts are normative). Videos were excluded if they were duplicates or had technical glitches (e.g., regular-length videos with no audio).

### Coding procedure

For each eligible video, we coded data about the source (uploader account name, account type, narrator characteristics) and modality (presentation style, e.g., animation, person talking on screen). Account type was determined by reviewing account names (e.g., BBC News would be identifiable as a news organization), and the “About” page, when necessary, applying predefined classification schemes for account type.

Our coding process was both *a priori* and emergent. While we began coding with our initial codebook, analysis of the transcripts revealed two additional relevant communication strategies*: use of storytelling* and *reference to additional learning resources*, which were added to the final codebook. To ensure reliability, three researchers jointly coded 15% of the transcripts, meeting weekly to discuss discrepancies and refine code definitions. This process repeated three times with different subsets of the data. For the final coding, two researchers independently analyzed all transcripts, holding weekly consensus meetings.

To assess comprehension, we analyzed transcripts using Readability Studio 2021 software. We calculated the Flesch-Kincaid score for readability ^35,36^ and Easy Listening Formula (ELF) scores for auditory comprehension ^37^. Both metrics provide grade-level estimates of comprehension difficulty.

### Analysis

To address our research questions, we employed a mixed-methods analytic approach. For RQ1 (communication strategies), we performed both quantitative and qualitative analyses. We first calculated the frequency of each coded strategy. Second, we qualitatively analyzed the coded content to understand how each corresponding strategy manifested, with two authors independently reviewing data to identify emergent sub-categories. We also conducted descriptive analyses of the readability and auditory comprehension metrics. For RQ2 (sources) and RQ3 (modalities), we conducted descriptive analyses of the metadata and manually collected characteristics.

## Results

We identified 294 YouTube videos that were eligible for inclusion in our study; 216 regular videos and 78 Shorts. All these videos were published between 2008 and 2023.

### RQ1: What communication strategies are used in YouTube videos about epigenetics?

Our analysis revealed how each communication strategy manifested and its explanatory purposes. The videos averaged Flesch-Kincaid readability scores of 10.7 and Easy Listening Formula (ELF) scores of 10.8 (both approximately 11th-grade level), indicating the content required higher than average comprehension skills (Table 2). All six codes represented distinct communication approaches, but they varied in their patterns of manifestation and in their association with specific epigenetic concepts. Here we describe how each communication strategy appeared in the videos and, where applicable, identify the epigenetic concepts they primarily helped to explain.

#### Use of metaphors, analogies, or similes ^2^

30.6% (90/294) of videos used metaphors, analogies, or similes, primarily to clarify the relationship between genes and epigenetics. Three dominant metaphorical patterns emerged. *Musical metaphors* portrayed genes as instruments and epigenetics as the musician. For example, “Your genes are like the keyboard on the piano, but the epigenome is the piano player controlling which genes are expressed” (transcript# 60). *Electrical metaphors* compared epigenetic regulation to light controls. For example, “Just like how a light switch can control the brightness of a room, epigenetic changes can control how genes are expressed” (transcript# 84). *Literary metaphors* framed DNA as text and epigenetic modifications as formatting. For example, “If … the genome is a book … Epigenetics influences gene function by changing the font size…” (transcript# 309).

These metaphors served three functions: First, they served to clarify biological processes, complex epigenetic mechanisms like DNA methylation (“… the methyl groups… are like sticky notes placed on the blueprint,” transcript# 277), and how environmental influences produced changes in gene expression (“Imagine your DNA as a script … and epigenetics as the director” transcript# 53). Second, metaphors served to illustrate the inheritance of trauma, health conditions, or behaviour. For example, “The sins of the fathers can be visited on the sons … by epigenetic inheritance” (transcript# 324). Third, metaphors served to help distinguish between genes and epigenetics by, for example, *c*ontrasting genetic determinism with epigenetics’ ability to be changed by personal action (“DNA is not destiny. You can write the very script of your life …” transcript# 142) or environmental influence (“Genetics loads the gun, but it’s the environment that pulls the trigger” transcript# 278).

#### Scientific examples

27.9 % (82/294) of videos used scientific examples, mostly focused on serving four explanatory functions: 1) demonstrating epigenetic inheritance, 2) illustrating environmental influences, 3) establishing disease risk and causation, and 4) explaining epigenetic aging. The most commonly mentioned examples were *historic case studies*, including the effects of famines in the Netherlands (1944-1945), Sweden (1867-1869), and Ireland (1845-1852). One video reported, “One example of epigenetic inheritance is the Dutch Hunger Winter … children … had a higher risk of obesity … due to epigenetic modifications” (transcript# 145). Other scientific examples were *laboratory experiments*, primarily focused on mouse models. As one video described, “… we experimented with resetting epigenetic structures in our aged mice, and we found that we could safely reverse blindness and rejuvenate kidneys and muscle” (transcript# 213). All but three scientific examples were studies based on animal models of epigenetics. Inheritance and environmental influences were explained through both historical and experimental examples, whereas disease mechanisms and aging were addressed exclusively through experimental findings.

#### Use of storytelling

22.1% (65/294) of videos told stories to explain epigenetic concepts. Five major story types emerged: First, personal stories, which told first-person accounts of individuals overcoming genetic predispositions, following a before-and-after narrative structure. For example, “I have rheumatoid arthritis in my genes … at 17 [before] … it’s been over 20 years now [after, by identifying the epigenetic causes and working with their body to change them], I don’t have rheumatoid arthritis anymore… it’s your epigenetics” (transcript# 118). Second, historic events which contained some of the scientific examples that were told like a story (e.g., a plot, having characters, and a setting). For example, “In the 1940s, pregnant women during the Dutch Hunger Winter famine gave birth to children who had higher rates of diseases … due to increased methylation …” (transcript# 111). Third, twin contrast where stories juxtaposed real or hypothetical identical twins to explain differential health outcomes. For example, “Let’s look at two identical twins … However, why is it that Armand develops a heart disease at the age of 40 while Strayon does not? It all has to do with epigenetics” (transcript# 140). Fourth, intergenerational trauma narratives where stories framed social injustices as a precursor to generational health disparities through social epigenetics — how social experiences can influence human health outcomes through epigenetic changes. For example, stories linking oppression to high rates of hypertension and diabetes in African-American communities, stating, “This trauma was embedded inside of the DNA, inside of the genomes” (transcript# 317). Others described healing rituals, “When we all gather to light the candles, we take a deep breath to release the week and greet the Sabbath. Afterwards, we kiss and hug, and sometimes even burst into song. We’re not just repeating traditions to keep them alive; we’re sealing them into our genes by imprinting a new code, one that gets passed along, one that lives in our body” (transcript# 328). Finally, biological allegories, where stories used known biological processes (e.g., metamorphosis) to explain epigenetic changes. For example, “The butterfly is a great example of epigenetics at work … When the larva becomes a caterpillar, its DNA doesn’t change; the expression of genes does … when it transforms into a beautiful flying butterfly, it still has the same DNA of a larva and caterpillar, but different genes express themselves” (transcript# 303).

#### Use of undefined jargon

21.8% (64/294) of videos used specialized terms without defining or explaining them (see Figure 1 for frequent undefined jargons). These undefined terms fell into four patterns: 1) *Core epigenetic processes* used epigenetic process-related terminologies without defining or describing them (e.g., “methylation,” or “histone modification”), 2) *Molecular biology terms* were used to describe genome-level biological processes other than epigenetic processes (e.g., “transcription,” “promoter,” “gene expression”), 3) *Field-specific acronyms* were often used without providing what the acronym stood for or what it meant (e.g., “HDAC4” [histone deacetylase] or DNMT1 [DNA methyltransferase]). 4) *Content-specific references* made unexplained allusions to concepts like “Yamanaka Factors” (transcript# 163) and were prevalent in videos discussing commercial or anti-aging research, where proprietary terms or novel mechanisms were introduced without sufficient contextualization.

**Figure 1.**
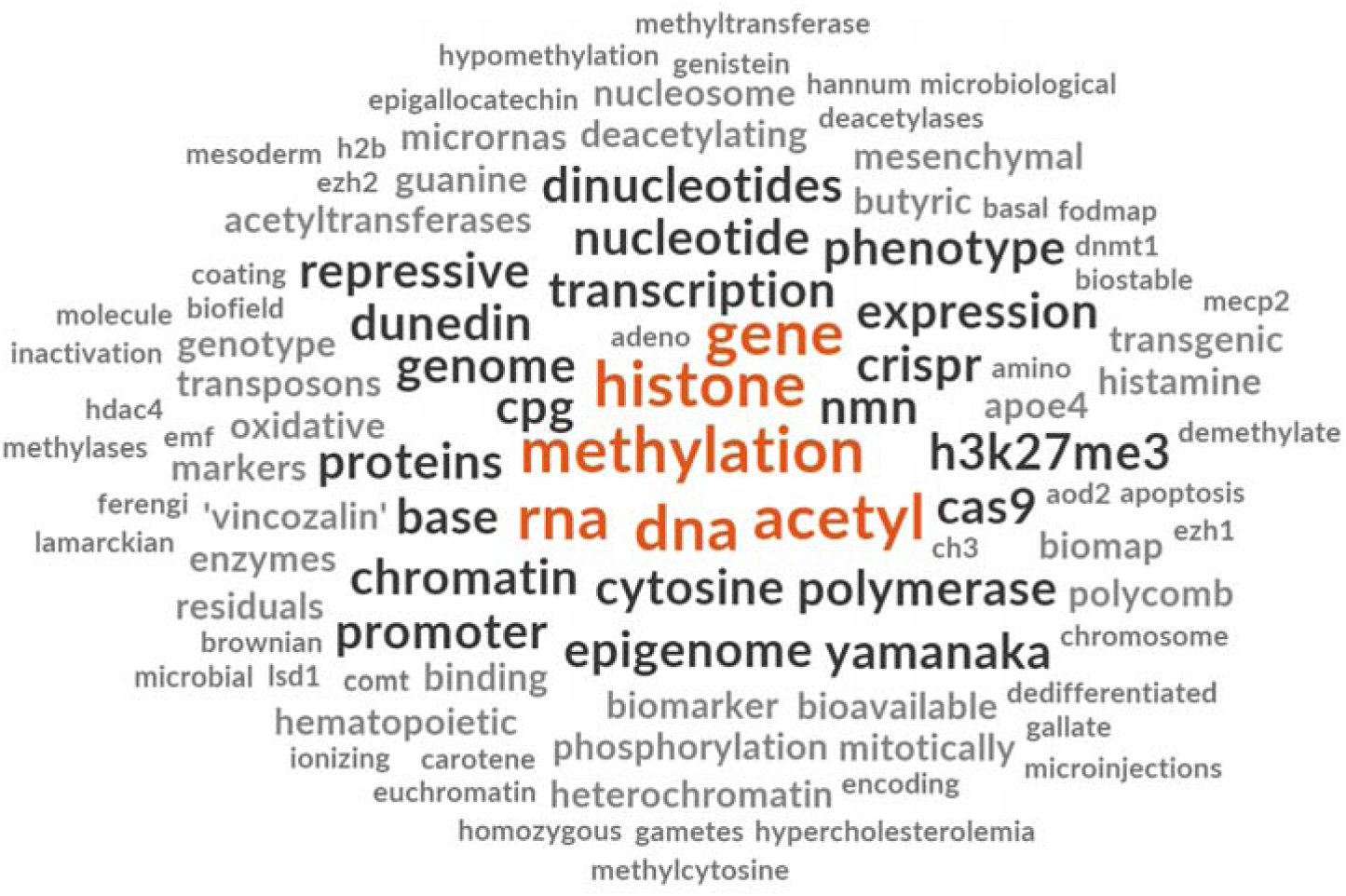
Word cloud of most frequently used undefined jargon in the videos.

#### Reference to additional learning resources

9.9% (29/294) of videos pointed to additional resources, typically at the video’s end, in four forms. *Academic and scientific resources* provided references to research institutions (e.g., “standupforcancer.org/clinicaltrials” transcript# 101; “Mayo clinic” transcript# 156; and “CDASH Genomics” transcript# 222). Notably, only one peer-reviewed paper (transcript# 310) and one university course (transcript# 234) were referenced as additional resources in the sample. *Commercial products* were also used as additional resources for audiences to learn more. These included books (e.g., A Poisoned Mind by Kurt Gassner, transcript# 154) or products like epigenetic testing kits (e.g., transcript# 325), and epigenetic age clocks (e.g., transcript# 94). *Expert consultations* named individuals such as “Bruce Lipton, Greg Barden, Hal Hills” as field experts (transcript# 172), encouraging audiences to look them up to learn more.

Finally, *digital resources* provided non-scientific/non-academic web sources such as creation.com (transcript# 339) or epigenetics.com (transcript# 278), and links to other YouTube content and creator guides.

#### Acknowledgment of scientific uncertainty

Only 7.5% (22/294) of videos acknowledged uncertainties regarding epigenetic concepts or discoveries. These acknowledgments appeared in three forms. Uncertainty about epigenetic discoveries and processes was expressed by *direct admissions of limited knowledge*. For example, “… scientists don’t even know yet exactly which switches the Swedish famines flipped … we’re a long way off from being able to make those connections in humans” (transcript# 174). Uncertainty about epigenetic topics was also expressed by framing *epigenetics as an emerging science*. For example, “Only recently have scientists started collecting evidence… It’s all part of an emerging field of science known as epigenetics” (transcript# 139). Another way of expressing uncertainty was the use *of tentative language*, such as modal verbs like may, might, or could. For example, “animal studies have suggested that … maybe also experiences of past generations can leave epigenetic memories” (transcript# 309).

Uncertainty was usually expressed regarding epigenetic inheritance in humans, environmental effects on epigenetic modifications, knowledge of epigenetic mechanisms, and disease causation by epigenetic modifications. However, no uncertainty was expressed regarding links between epigenetics and individual behaviour, stress, or trauma.

### RQ2: What are the sources disseminating epigenetic information on YouTube?

The epigenetic videos in Table 1 predominantly originated from independent organizations (53.4%, e.g., SciShow, Veritasium) that exclusively create online content and post videos on YouTube. This type of source was followed by personal accounts of independent online content creators (26.9%); these were different from production-company-style online content creators of independent organizations. Other account types, including educational institutions (3.1%), healthcare industries (3.7%), and news organizations (2.7%) were far less common. Presenters were most often male-presenting (34.7%) or female-presenting (22.1%), with a small subset featuring presenters of each gender presentation (4.4%). A large proportion (35.7%) of videos lacked any visual indication of the possible sex of the narrator. Racial presentation was more frequently white-presenting speakers (44.9%) than non-white-presenting speakers (13.3%). A substantial proportion of videos (37.1%) lacked any visual indication of the possible race of the narrator. Among only the videos *with* any visual narrator, 56.7% were male-presenting and 73.7% were white-presenting.

**Table 1:**
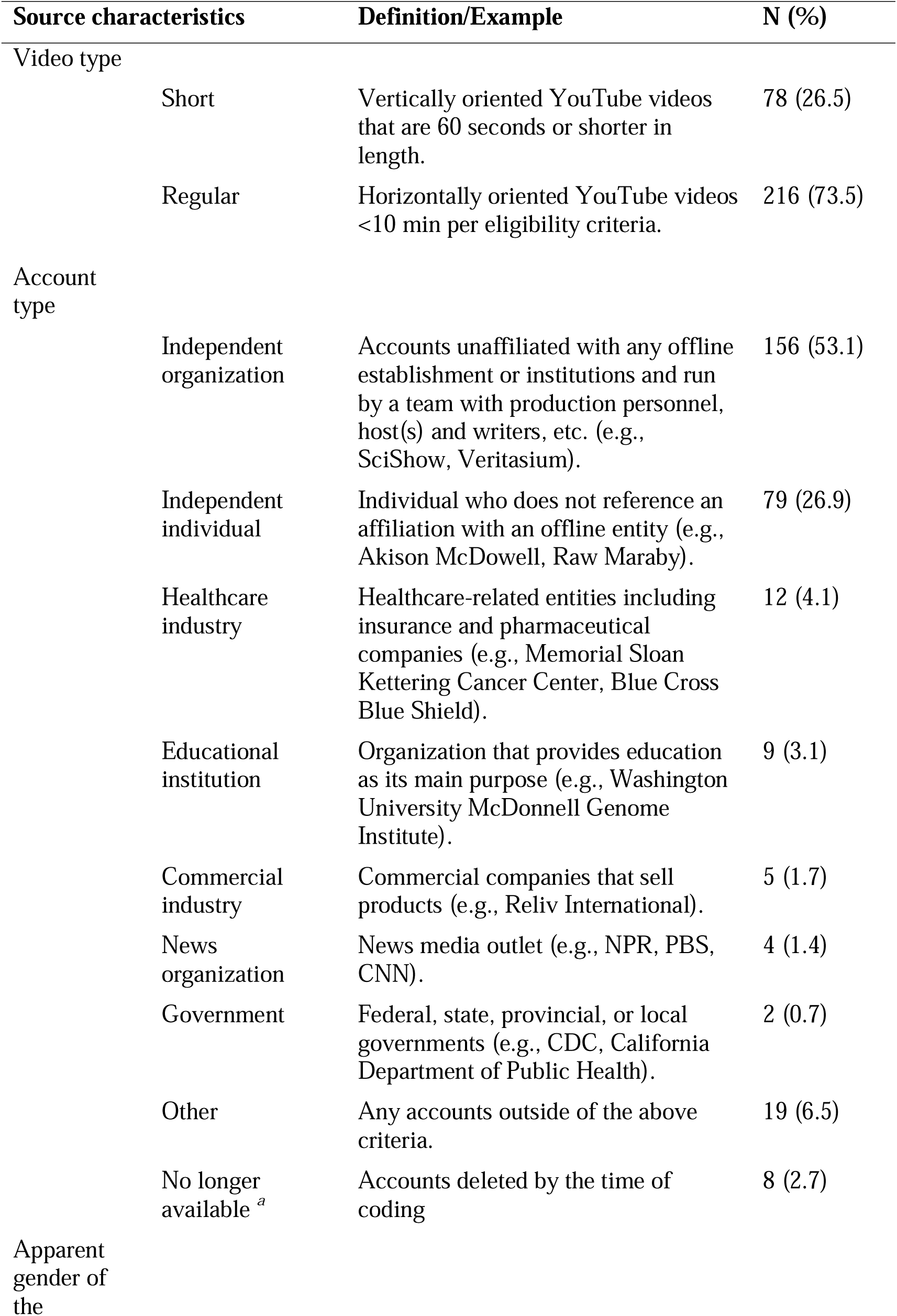
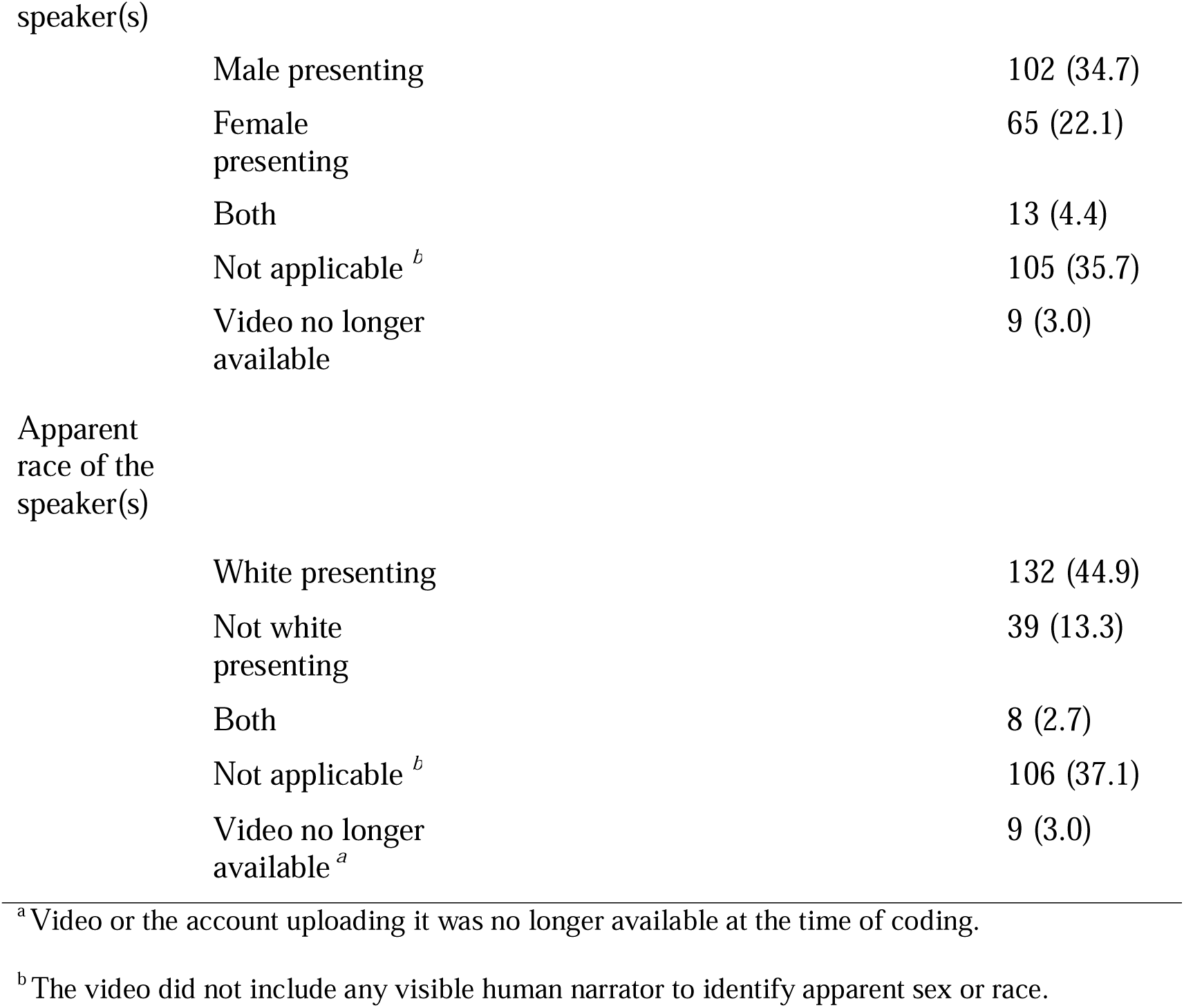
Source characteristics (*N* = 294 YouTube videos).

### RQ3: What modalities are used in YouTube videos to communicate epigenetic information?

The analysis of video modalities (Table 2) revealed that the most common presentation modality was a person talking on screen (40.1%), followed by animation and/or graphics with voiceover (37.4%). Other modalities included slideshows with voiceover (16.0%), conversations between multiple people (16.0%), and text on graphics without voiceover (4.4%). Fifty-one (17.4%) videos used multiple modalities in their presentation style and were double coded.

**Table 2:**
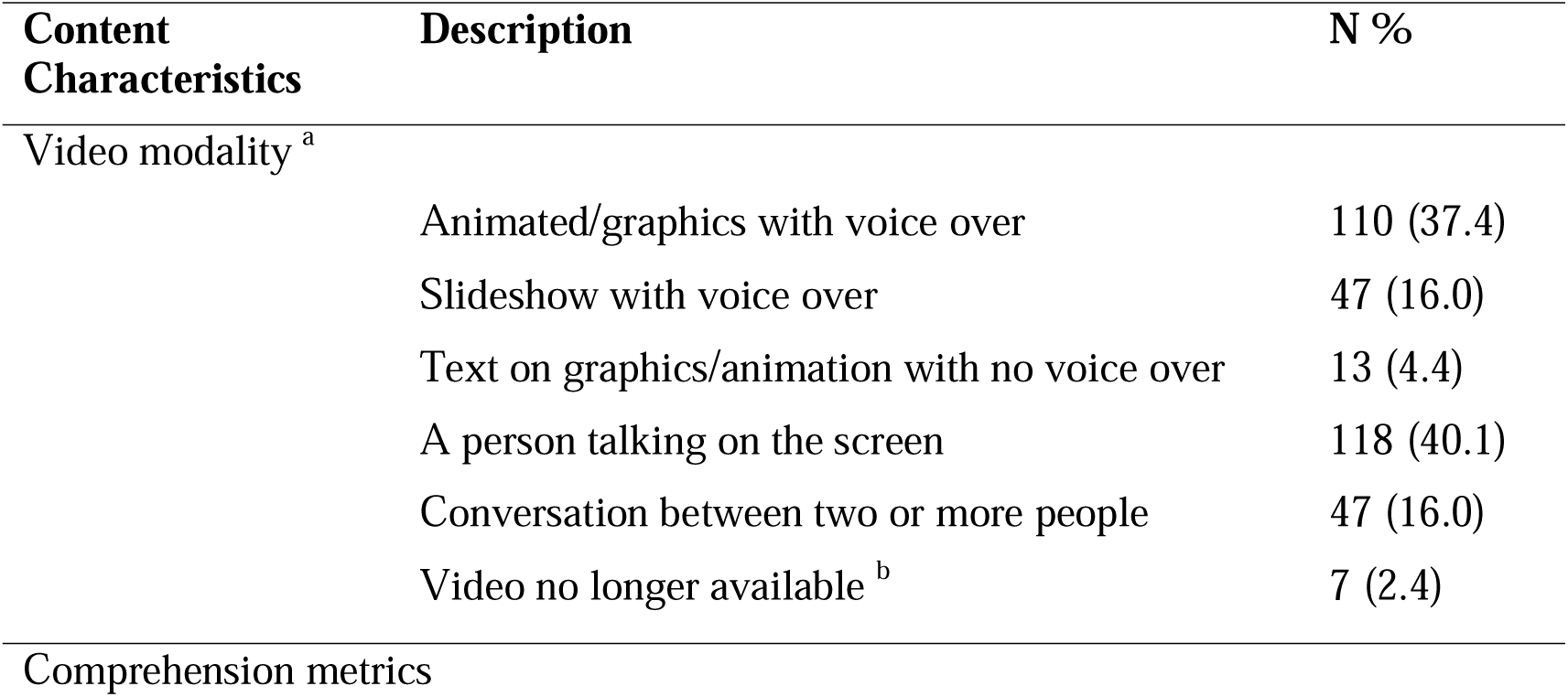
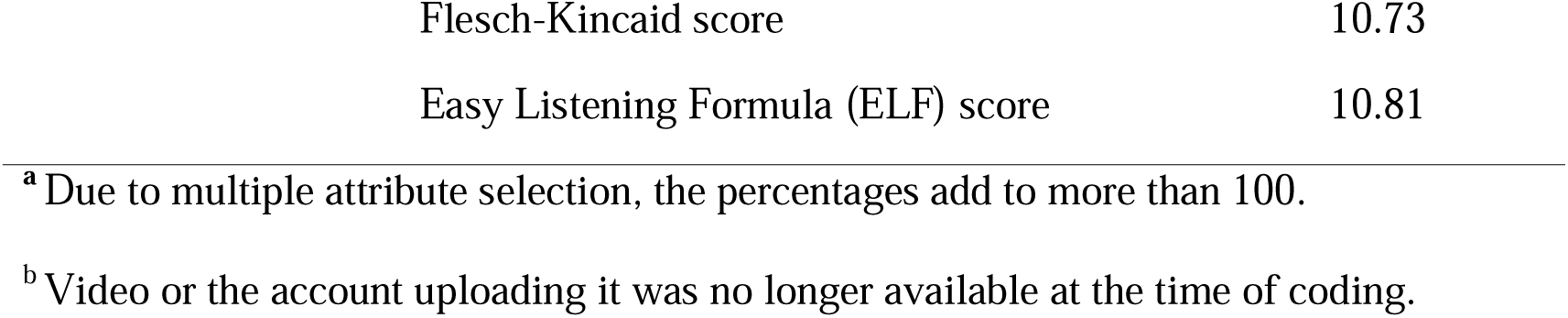
Content characteristics (*N* = 294 YouTube videos).

| **Content Characteristics** | **Description** | **N %** |
| --- | --- | --- |
| Video modality <sup>a</sup> | Animated/graphics with voice over | 110 (37.4) |
|  | Slideshow with voice over | 47 (16.0) |
|  | Text on graphics/animation with no voice over | 13 (4.4) |
|  | A person talking on the screen | 118 (40.1) |
|  | Conversation between two or more people | 47 (16.0) |
|  | Video no longer available <sup>b</sup> | 7 (2.4) |
| Comprehension metrics |  |  |

|  |  |
| --- | --- |
| Flesch-Kincaid score | 10.73 |
| Easy Listening Formula (ELF) score | 10.81 |

<sup>a</sup> Due to multiple attribute selection, the percentages add to more than 100.
<sup>b</sup> Video or the account uploading it was no longer available at the time of coding.

## Discussion

Our analysis of epigenetic content on YouTube revealed three key patterns. First, we identified significant limitations in the application of science communication best practices. Second, we found that these videos were predominantly published by independent organizations and individuals that were unaffiliated with any formal scientific or health institutions. Finally, when visible narrators were present, they were more likely to be male-presenting than female-presenting and more frequently white-presenting than other racial identities.

### Gaps in science communication best practices

Our comparison of epigenetics videos with science communication best practices revealed several shortcomings. For example, metaphors, analogies, and similes (termed as metaphors henceforth), which can be used to clarify complex ideas ^40–47^, and explain abstract concepts ^48^, were used in fewer than one-third of the videos. When present, we found that metaphors were often used to contrast genetics (framed as static, passive, or immutable) with epigenetics (framed as dynamic, active, or malleable). While phrases like “genes are not destiny” can counter genetic determinism, metaphors such as “the environment loads the gun” risk reinforcing environmental determinism—the idea that exposure to certain environmental factors “destines” individuals to negative outcomes ^49–51^. This messaging can be harmful, particularly for children and adults affected by famine, war, or genocide, as it may imply that those living outside Western “standards” are lesser ^52–54^. Additionally, metaphors portraying epigenetics as a person (e.g., “piano player” or “director”) “controlling” an object (e.g., “piano” or “play”) may wrongly suggest individuals have excessive control over their epigenome, which could contribute to victim-blaming and stigma. Future research is needed to investigate the effect of using such metaphors on public conceptualization and perceptions of epigenetics.

Providing concrete examples is another way to enhance comprehension of abstract topics like epigenetic processes ^55,56^, but only a quarter of our sample used this strategy to explain how epigenetics works. Those who used examples often focused on European case studies, reflecting a Eurocentric bias in science, especially on generational effects of famine — which mostly have occurred outside Europe ^57^. This perpetuates a broader Eurocentric bias in genetic sciences ^58,59^.

Storytelling is another powerful persuasive tool because it can “transport” audiences into a narrative, whether factual or fictional ^60^. While often viewed negatively in science as “anecdotal evidence,” studies show storytelling promotes narrative processing, more effective than pragmatic processing for explaining science ^61^. Yet, less than a quarter of videos in our study used storytelling for epigenetics and related concepts.

Field-specific terms should always be explained in science and public communication, as undefined jargon can hinder comprehension ^62^. Unexplained jargon in intergroup communication (e.g., between science communicators and public audiences) reduces credibility ^63^ and increases intergroup distance, leading to negative perceptions like seeing the YouTubers as outgroup or paternalistic ^64,65^. Although most videos in this study defined jargon, over a fifth did not.

Expressing uncertainty in science communication can increase credibility ^66,67^ but it can also backfire. For example, studies on vaccine safety found that uncertainty can exacerbate hesitancy ^68^. A systematic review offered guidance on the effects of uncertainty in science communication, categorizing them by type ^69^. The review found that uncertainty stemming from scientific disagreement or competing paradigms consistently has negative effects on audience trust. None of the videos in our sample included this form of uncertainty. Instead, the videos primarily expressed uncertainty in terms of limited knowledge or epigenetics being an emerging field which has mixed effects on audience trust and credibility ^69^. The tentative language used in our sample aligns with the review’s “scientific uncertainty” category, referring to the inherent lack of absoluteness in scientific findings. However, this type of uncertainty has also produced positive, negative, and null effects on audience trust and credibility ^69^. In our study, over 90% of the videos failed to acknowledge the uncertainty surrounding epigenetic scientific findings. While this omission may not directly impair the explanation of concepts, excluding expressions of uncertainty risks leading the public to misunderstand and overstate the implications of epigenetic discoveries ^39^, despite the mixed effects on trust and credibility.

Our finding that video transcripts had higher comprehension difficulty suggests the content was limited to audiences with at least some high school education. In the United States, 71% of the population has completed high school education, while globally, this figure goes down to 61% ^70^. Moreover, the average American reads at a 7^th^ to 8^th^ grade level ^71^. A key priority for epigenetic communicators on social media should be the use of strategies that enhance public comprehension of science ^72,73^.

Finally, videos that cited experts often referenced individuals who were primarily proponents of alternative medicine rather than specialists in epigenetics. This could lead to misconceptions about the field’s scope, capabilities, and limitations. We found only one peer-reviewed article as a resource recommended for further learning, compared with multiple references to commercial products and popular press books. This finding underscores the need to enhance the accessibility of peer-reviewed literature. Doing so would provide non-expert audiences with credible learning resources, reducing their reliance on non-expert materials based solely on comprehensibility ^74^.

### Lack of expert sources and demographically inclusive narrators

Institutional sources such as the healthcare industry, educational institutions, and news organizations were notably underrepresented in the sample. This distribution contrasts with other areas of science communication on YouTube. For example, COVID-19 vaccine videos from news agencies and medical institutions made up about 40% ^75^, while healthcare industry videos accounted for nearly two-thirds in spinal cord stimulation content ^76^. Additionally, universities were the main publishers for Alzheimer’s disease videos ^77^. Notably, no videos from professional genetics or medical societies, such as the Genetics Society of America or the American Society of Human Genetics, appeared in our sample. The absence of field experts on YouTube or other social media platforms, while there is abundant content on epigenetics, implies that non-experts and science influencers provide the bulk of the information on epigenetics.

Together with the fact that additional sources provided in most videos are non-academic or non-scientific, this absence of experts can lead to exaggerated claims about epigenetics, as seen in the content analysis of topics about epigenetic videos on YouTube ^19^). The future of science communication may be on social media platforms ^78^. If scientists and professional organizations are not leading the dissemination of scientific discoveries in spaces where most of the public is consuming scientific information, that void is filled by non-experts, dubiously funded influencers, and communicators spreading misinformation and disinformation ^79,80^.

Video narrators who were visible were often male-presenting and white-presenting; this reflects known inequities in science communication ^81–83^ and is suggestive of the “Matilda effect” whereby male presenters are viewed as more credible ^84,85^. Although research on YouTube science communicators’ demographics remains limited, existing studies report similar patterns, with one analysis finding 76.9% of science communicators in their sample were male ^86^. The existing literature emphasizes the importance of representation, showing that race and gender concordance can enhance understanding, interest, and trust ^87^. It remains unclear if videos without visible narrators involve individuals of other genders or racial backgrounds. Future studies should explore how perceptions of gender and race affect content creators’ choices of metaphors, scientific examples, framing of scientific concepts, personal responsibility narratives, and other messaging in educational YouTube videos.

### Limitations and future directions

Our study’s algorithmic search constraints may limit generalizability to all YouTube videos on epigenetics, especially those longer than 10 minutes. Future work should stratify by video purpose (e.g., educational vs. commercial) to identify context-specific communication patterns. The immediate next step for the current study would be to develop creator guidelines for epigenetic communication on YouTube and other visual social media platforms. Future research could explore audiences’ understanding of epigenetics through the metaphors used in these videos to inform future public education efforts.

## Conclusion

Together with our prior findings on exaggerated claims in YouTube videos about epigenetics, the present analysis reveals a two-dimensional challenge: epigenetic communication on YouTube is compromised by both *what* is said and *how* it is said. Specifically, the current study identifies systemic issues that help explain why such exaggeration exists, namely, underutilization of best practices in scientific communication, predominance of creators unaffiliated with medical or scientific institutions, and narrow distribution of demographic characteristics among presenters. Greater active presence of professional science communicators from expert organizations could mitigate the spread of exaggerated claims. While metaphors and examples aid understanding, their deterministic or Eurocentric framing risks perpetuating exclusionary science communication and alienating audiences from demographically diverse backgrounds. Prioritizing inclusive, accessible, and transparent messaging is critical for this evolving field.

## Funding Information

Grant Number: **R01ES033743** (PI: Erika A. Waters)

Funder: **National Institute of Environmental Health Sciences**

Grant Number: **P30 CA008748** (Awardee: Jada G. Hamilton)

Funder: **National Cancer Institute**

Grant Number: **T32CA190194** (Awardee: Aantaki Raisa)

Funder: **National Institutes of Health**

## Ethical Approval and informed consent statement

Since the study was conducted by collecting publicly available data on YouTube, no ethical approval was sought or needed.

## Data availability statement

Video transcripts and the codebook can be found on the Open Source Framework platform by clicking here: https://osf.io/rft9s/overview?view_only=144bc8bb9c9f487a97f8e8ced68cdcf5

## Conflict of Interest

The authors declared no potential conflicts of interest with respect to the research, authorship, and/or publication of this article.

## Notes

### Competing Interest Statement

The authors have declared no competing interest.

